# Rapid diagnosis of fever etiology using wearable temperature monitoring and machine learning

**DOI:** 10.64898/2026.07.27.26359010

**Authors:** Shihan N. Khan, Seungwoo Lee, Xiheng Ren, Emily Wittrup, Rashmi Madhukar, Christopher Flora, Kelly Mayhew, Michelle Rozwadowski, Eric Winnega, Kay Leopold, Jason B. Weinberg, Jonas Paludo, Adam F. Binder, Monalisa Ghosh, David Frame, Erin Craig, Thomas M. Braun, Rishi Chanderraj, Anthony D. Sung, Kayvan Najarian, Sung Won Choi, Muneesh Tewari

## Abstract

**Introduction:** Distinct temperature patterns have long been recognized to correlate with fevers of differing etiologies. While the use of wearable sensors for high-frequency temperature monitoring (HFTM) on a near minute-by-minute basis has been shown to detect fevers earlier than standard-of-care nursing vital sign assessments in hospitalized patients, leveraging these high-resolution datasets to computationally identify unique digital signatures for real-time diagnosis of underlying fever etiology has not been widely explored. Diagnostic uncertainty is common in patients undergoing hematopoietic stem cell transplantation (HCT), with only 20–30% of febrile neutropenic episodes being microbiologically documented. We hypothesized that unique temperature patterns extracted from HFTM data collected during episodes of febrile neutropenia could be used to develop a supervised machine learning classifier capable of accurately predicting underlying fever etiology in HCT patients.

**Methods:** We analyzed 68 clinically independent fever episodes recorded in HCT patients (n=90) outfitted with an FDA-cleared wireless temperature sensor (TempTraq®, BlueSpark Technologies) that measured axillary temperature every 2 minutes throughout hospitalization. Time-series features were extracted from temperature traces spanning 1 hour before to 3 hours after fever onset and used to train a suite of machine-learning models to distinguish engraftment fevers from other fever etiologies. Model training and evaluation were performed using repeated stratified 5-fold patient-level cross-validation, yielding 100 train-test evaluations.

**Results:** Among all classification models, the logistic regression classifier provided the best overall performance and interpretability, achieving 94% specificity (95% CI, 0.84–1.0) for identifying engraftment fevers with a mean AUROC of 0.88 ± 0.10. Feature importance analysis demonstrated that both clinical variables and HFTM-derived temperature dynamics contributed to model performance, with a strong reliance on time-series features captured within the first 4 hours of fever onset.

**Conclusion:** Our study provides a demonstration that continuous temperature data collected from patients outfitted with wearable sensors can be leveraged not only for early fever detection but also for machine learning–based diagnosis of fever etiology. These findings suggest that dynamic temperature patterns contain clinically meaningful physiologic information that with further studies could support real-time diagnostic decision-making and guide safe de-escalation of empiric antibiotics during febrile neutropenia in patients undergoing intensive cancer therapy.

---

Due to the high morbidity, mortality, and economic burden associated with infection-related febrile neutropenia (FN), empiric broad-spectrum antibiotic therapy remains the cornerstone of initial management even in the absence of a documented source of infection. Current guidelines from the Infectious Diseases Society of America (IDSA) and the European Conference on Infections in Leukemia (ECIL) recommend administering intravenous antibiotics within sixty minutes of fever onset in high-risk neutropenic patients, such as those undergoing hematopoietic stem cell transplantation (HCT)^1–3^. While potentially lifesaving in some cases, studies show that a causative pathogen is identified in only 20–30% of FN episodes ^4–6^, and as few as 5–10% result in serious bacterial infections^6,7^. This prescribing practice has downstream consequences, as unnecessary antibiotic use drives antibiotic resistance. Furthermore, multiple studies demonstrate that broad-spectrum antibiotic use is associated with increased risk and severity of acute graft-versus-host disease following HCT^8–10^, suggesting that the use of empiric antibiotics when not needed is not benign in this patient population that is already at high-risk for therapy-related complications. Additionally, the cost of FN-related hospitalizations in the United States has been estimated at over $25,000 per episode, driven largely by empiric broad-spectrum antibiotic use, prolonged hospital stays, and supportive care requirements^11,12^. As such, there is an ongoing need to develop better clinical tools for distinguishing true bacterial infections requiring antibiotics from other causes, which can guide antibiotic stewardship to improve patient outcomes.

The integration of mobile health technology into cancer care is rapidly transforming the management of cancer patients, offering novel tools for disease surveillance and early clinical intervention. Continuous vital signs monitoring on a near minute-by-minute basis—enabled by wearable, non-invasive sensors—has been shown to detect fevers earlier than standard-of-care (i.e., intermittent temperature measurement by hospital staff), in which vital signs are typically measured every four to eight hours in the hospital^13^. While prompt identification of fever is important, early detection alone does not resolve the challenge of accurately distinguishing those episodes of FN that are due to bacterial infections, and therefore need antibiotics, from fevers due to other causes, which do not need antibiotic treatment. In the setting of HCT, due to diagnostic uncertainty at the time when fevers are first detected, even fevers that are suspected to be non-infection, such as those triggered by hematopoietic stem cell engraftment (i.e., “engraftment fevers”), are routinely treated with empiric antibiotics in order to avoid missing a true infection. Thus, the ability to provide early, accurate diagnosis of the underlying etiology of a fever—i.e distinguishing bacterial infection from other processes—would be of great clinical importance. Building on our group’s previous work utilizing high-frequency temperature monitoring (HFTM) for early fever detection in HCT patients^13,14^, we hypothesized that the *dynamics* of temperature patterns measured at high time-resolution may contain unique digital signatures diagnostic of specific fever etiologies. We therefore sought to apply machine learning techniques to the dynamic temperature profiles of fever events captured in HCT patients wearing a continuous temperature sensor, in order to identify unique patterns differentiating bacterial infection-related versus other fever etiologies (e.g., non-infection engraftment fevers, etc.).

To explore this idea, we conducted a prospective observational study of HCT patients outfitted with an FDA-cleared, non-invasive temperature sensor (TempTraq®, BlueSpark Technologies) applied to the axilla as an adhesive patch that continuously measures body temperature every two minutes in parallel to standard-of-care (SOC) nursing vitals (i.e., every four to eight hour temperature measurements) during their respective hospitalizations (Fig. 1a, blue panel). After providing IRB-approved informed consent, patients enrolled in the study were asked to *self-apply* HFTM patches to be worn daily and replaced every 24 to 72 hours, depending on patch version, for as many days of the hospitalization as was convenient for each study participant. HFTM datasets underwent pre-processing (Fig. 1a, orange panel) in which date and time stamps were standardized and masked as ‘days post-infusion’ relative to each subject’s transplantation date. Of the patients with at least 12 hours of cumulative HFTM data recorded during their hospitalization (n=90), HFTM captured 45-fold more temperature readings (n= 833,544) than nursing SOC vitals (n= 18,700) during the same time period, representing a median data capture of 126 hours per patient. Both HFTM and SOC temperature data were then systematically screened for fevers, defined as ≥3 independent HFTM readings above 38°C within a 60-minute window or a single SOC temperature measurement greater than 38°C. Across the study cohort 47 patients experienced at least one fever episode, with several of these study subjects having multiple febrile events during their hospital course, resulting in a total of 85 discrete fever events detected during the study period with concurrent HFTM and SOC data available for analysis (Supp. Fig. 1).

**Figure 1.**
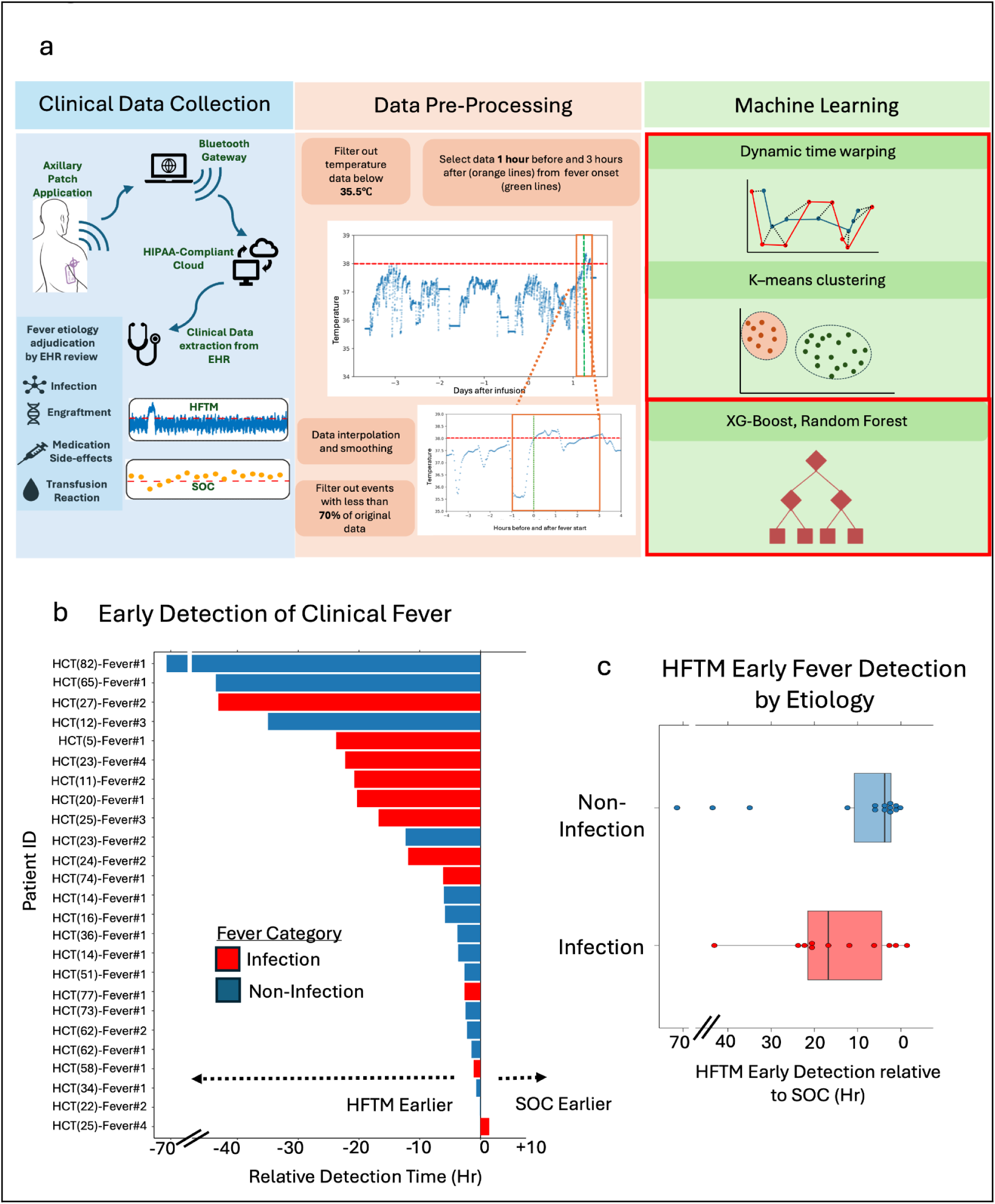
(a) Overview of methodology for clinical temperature data capture and machine learning analysis. HFTM data collected from patients via TempTraq® was transmitted to a HIPAA-compliant cloud through a Bluetooth-connection based gateway device. Subject demographics, nursing SOC temperature data, dates of admission, stem-cell transplantation, and discharge from hospital were obtained from the electronic health record (EHR) and used by physician reviewers to adjudicate fever etiology as infection, engraftment, medication side-effects, or transfusion reaction for each fever event (blue panel). Data pre-processing included filtering out temperature readings below 35.5°C, selecting data within 1 hour before and 3 hours after fever onset, interpolating and smoothing time-series measurements, and excluding events with less than 70% original data coverage (orange panel). The processed data was subsequently analyzed using machine learning approaches, including dynamic time warping and K-means clustering as an initial analysis, and subsequently using a suite of classification models such as logistic regression, random forest, amongst others (green panel). **(b) Early detection of clinical fever by high-frequency temperature monitoring (HFTM) compared to standard-of-care (SOC) intermittent nursing assessments.** Relative detection times for individual patients are shown, categorized by fever etiology (infection [solid red], and non-infection [blue] causes). HFTM detected 24 out of 25 concordant fevers earlier than SOC, and these were detected a median of 6.1 hours earlier. Negative time values indicate earlier detection by HFTM relative to SOC. **(c) Early fever detection by HFTM stratified by infection versus non-infection etiology.** Box plots show the distribution of lead times for fever detection by HFTM relative to SOC, stratified by infection (red) and non-infection (blue) etiologies. In fevers adjudicated as infection-related, HFTM achieved a median lead time of 18.6 hours prior to SOC documentation. For non-infection fevers, HFTM identified events a median of 3.7 hours earlier than SOC.

First, we assessed agreement in fever detection between HFTM and SOC vitals by performing a concordance analysis (see Supplemental Methods), classifying detected fevers into three groups: (a) concordant fevers, detected by both SOC and HFTM data streams within a 24-hour window; (b) discordant HFTM-only fevers, detected exclusively in the HFTM data stream without a corresponding SOC fever within 24 hours; and (c) discordant SOC-only fevers, captured only in SOC but missed by HFTM within 24 hours. This concordance analysis revealed that 50 fevers were ‘concordant’, detected by both HFTM and SOC monitoring within an overlapping 24-hour window. When these concordant fevers were then retrospectively annotated by a team of clinicians to determine their etiology based on review of the electronic medical record (EMR) and using a systematic etiology annotation approach (see Supplemental Methods, Supp. Fig. 2), we found that 18 out of these 50 fevers were associated with detection of a specific pathogen (i.e. bacterial, viral, or fungal). Another 6 concordant fevers were associated with episodes in which clinicians had sufficient suspicion for bacterial infection to initiate antibiotics, but no pathogen was identified on culture (i.e., culture-negative FN). However, the majority of concordant fevers were annotated as ‘non-infection,’ with 15 fevers attributed to engraftment, and 11 fevers attributed to ‘other’ etiologies including medication side-effects, transfusion reactions, or not attributable to any clear etiology (Supp. Table 1).

Next, we evaluated the initial timing of concordant fevers detected in patients who had at least one fever event during their hospitalization (Supp. Fig. 3). We found that HFTM identified 24 out of 25 concordant fevers a median of 6.1 hours earlier than intermittent SOC nursing assessments (Fig. 1b). Notably, in fevers adjudicated as infection-related (n=11), HFTM achieved an even greater lead time, with detection occurring a median of 18.6 hours prior to SOC fever detection (Fig. 1c). For non-infection fevers (n=14), earlier detection by HFTM was still observed, with a median of 3.7 hours in advance of SOC (Fig. 1c). These findings corroborate the findings of studies demonstrating early fever detection by wearable continuous temperature monitoring devices, underscoring its potential to enable more timely clinical interventions in the correct clinical setting^13–15^.

We then investigated discordant fever episodes, first examining the 35 discordant fever episodes identified by HFTM (i.e., “HFTM-only”) but not detected by intermittent SOC monitoring (Supp. Fig. 1). Of these HFTM-only events, retrospective clinical adjudication indicated that 9 events were associated with infections requiring antibiotics in the subsequent seventy-two hours, whereas the majority—26 episodes—were deemed by clinician adjudication as non-infection, most often benign engraftment-related fevers (n=19). Reviewing the 20 discordant “SOC-only” fevers that were not detected by HFTM, we found that 19 of these 20 fevers simply did not have sufficient available coincident HFTM data (i.e., >80% missing or poor quality HFTM data - see Supplementary Methods for details), related in many cases to device connectivity issues for early participants in the study that were later resolved by our study team (Supp. Fig. S1). The one additional fever not formally captured by HFTM did in fact show multiple >38°C readings in HFTM data prior to SOC, but they did not meet our stringent criteria of ≥3 independent temperature readings above 38°C within a 60-minute window, and thus was classified as ‘not detected’ by HFTM. Taken together, these findings suggest that when connectivity is intact with properly applied sensors, HFTM rarely misses a fever event compared to SOC monitoring.

We then proceeded to address the key objective of this study, which was to investigate whether it is possible to use HFTM data collected using wearable devices to not only detect fever earlier, but also to accurately “diagnose” fever etiology in near ‘real-time’ (i.e. within several hours of the start of fever). Given that distinctive temperature patterns have long been noted by clinicians to correlate with fevers of certain origins (e.g., malaria, lymphoma etc.), we hypothesized that computational analysis of HFTM data could be used to identify unique, dynamic digital signatures associated with different underlying fever etiologies, whether infection or non-infection.

To explore the validity of this hypothesis, we first used a “pattern discovery” approach to investigate potential associations between dynamic temperature patterns and fever etiology using HFTM data (Fig. 1a, green panel). To do so, we applied unsupervised *k*-means clustering with dynamic time warping (DTW) to available HFTM data extracted from a time window spanning 1 hour before and 3 hours after onset of the first concordant fever (see Supplementary Methods and Supp. Fig. 4), to identify different types of dynamic temperature patterns present across patients (i.e., through a clustering approach). Based upon an analysis using the elbow method^16,17^, we set the number of clusters at three (Supp. Fig. 5). HFTM data in different assigned clusters were visualized with time series plots, depicting individual fever events in grey and the corresponding cluster centroids in blue (Fig. 2a), alongside bar graphs representing the distribution of infection-associated and non-infection fever events within each group (Fig. 2b).

**Figure 2.**
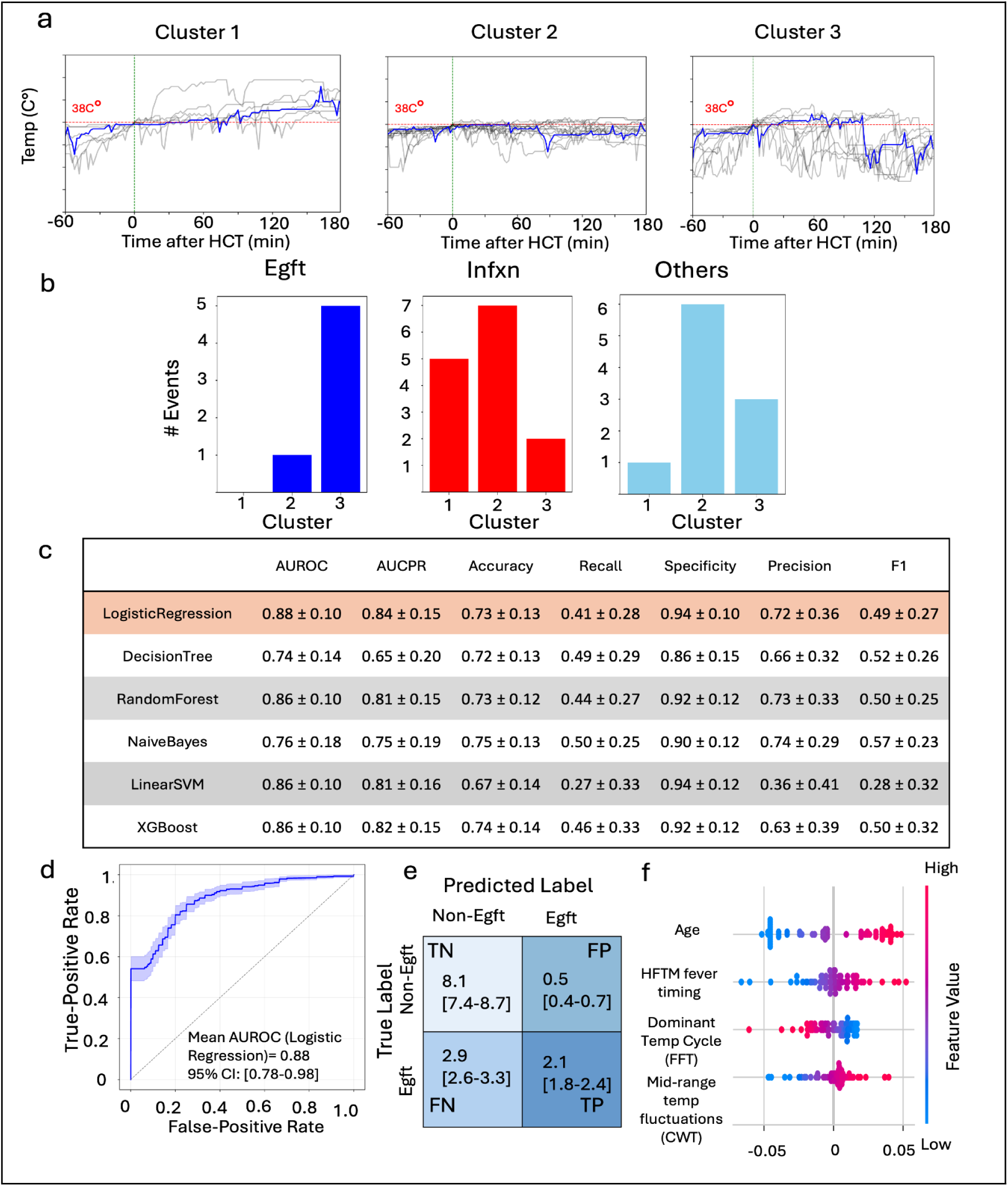
Clustering and classification of high-frequency temperature monitoring (HFTM) data for fever etiology discrimination. (a-b) K-means clustering with dynamic time warping (DTW) was applied to all available HFTM data spanning 1 hour before and 3 hours after onset of concordant fever events (n=30). Time series plots display individual fever events (grey) and cluster centroids (blue) for each cluster. Bar graphs depict the distribution of adjudicated etiologies—engraftment (blue), infection (red), and other non-infection causes (light blue)—across the three identified clusters. **(c) Performance metrics of various machine learning classifiers** (logistic regression, decision tree, random forest, naive Bayes, linear SVM, and XGBoost) for fever etiology discrimination. **(d) Receiver operating characteristic (ROC) curve for the logistic regression classifier.** The area under the mean ROC curve (AUROC) was 0.88 (95% CI: 0.73–0.95) and is represented by the dark blue line plot, estimated across 100 iterations of patient-stratified 5-fold cross-validation (20 repeats). The lighter blue shaded region represents the 95% confidence interval (CI) around the mean ROC curve. (**e**) **Average confusion matrix for the logistic regression engraftment classifier across 100 test-set iterations (5-fold cross-validation with 20 repeats).** Egft, engraftment fever; Non-egft, non-engraftment fever; TP, true positive; TN, true negative; FN, false negative; FP, false positive. **(f) SHAP summary (beeswarm) plot of feature contributions to the logistic regression model predicting engraftment.** Each point represents one of the [n = 68] fevers in the test set. Features are ordered top-to-bottom by mean absolute SHAP value (decreasing global importance). The horizontal position of each point gives that feature’s SHAP value — its signed contribution to the model’s output for that individual, in log-odds; points to the right of the zero line push the prediction toward higher likelihood of the engraftment (class 1), points to the left toward non-engraftment (class 0). Point color encodes the feature value (blue = low, pink/red = high), so the color–position relationship indicates the direction of each feature’s effect.

This clustering analysis revealed distinctive patterns of temperature dynamics that showed some preponderance based on etiology of fever, most notably for “engraftment” fevers which preferentially clustered within Clusters 2 and 3, with the majority being found in Cluster 3 (Fig. 2b, left; dark blue). In contrast, fevers associated with infections (Fig. 2b, middle; red) appeared more heterogeneous, spanning all three clusters (Fig. 2b, right; light blue).

Fever occurring around the time of bone marrow engraftment, commonly defined as the first of three consecutive days of achieving a sustained peripheral blood neutrophil count greater than 500 x10^6^ /L^18^, is a frequent phenomenon in the post-transplant period, reported in 10-77% of patients depending on the population, with a recent study demonstrating an incidence of 36% among 124 patients undergoing autologous HCT^19–21^. While severe engraftment syndrome, characterized by fever in combination with organ dysfunction such as pulmonary edema, can represent a serious and potentially life-threatening issue, the majority of HCT patients experience only transient, non-infection fever related to engraftment, which resolves with supportive care alone without the use of antibiotics. However, given that patients are typically neutropenic when ‘engraftment fevers’ develop, broad-spectrum antibiotics are commonly initiated per standard neutropenic fever protocols, as bacterial infection often cannot be easily ruled out by treating clinicians. Specific and reliable identification of fever patterns differentiating benign engraftment fevers from infection could therefore potentially reduce unnecessary use of empiric antibiotic therapy during the peri-engraftment period.

Given that engraftment-related fevers appeared to follow a distinct temperature pattern in our clustering analysis (Fig. 2a, 2b), we next sought to train a supervised machine-learning classifier using time-series features extracted from HFTM data along with basic clinico-demographic information, with the goal of differentiating engraftment fevers from other fever etiologies using HFTM data proximate in time to fever detection (i.e., from 1 hour before to 3 hours after the fever onset). While our present work is a proof-of-concept study, we incorporated the clinical motivation to create a model optimized for high specificity, which would ultimately allow clinicians to confidently identify engraftment fevers and safely de-escalate empiric broad-spectrum antibiotics during neutropenia in a subset of patients, while also minimizing false positives that could risk missing true infections in this high-risk population.

For ML analysis, we included both concordant and HFTM-only fevers. Multiple fever events from the same patient were permitted when subsequent fevers within the same inpatient course satisfied prespecified criteria for clinical independence (see Supplementary Methods). This yielded a total of 68 fever episodes that were used for ML analysis. To capture a range of dynamic fever features for model training, we first extracted 621 time-series features from HFTM fever traces after filtering (see Supplementary Methods and Supp. Fig. S6) using the *tsfresh* Python package^22^, quantifying statistical, temporal, and frequency-based properties of each temperature signal across the same time window used for clustering analysis, spanning one hour before to three hours after fever onset (see Supplementary Methods). Feature selection was performed using the Boruta algorithm^23^ to identify informative variables from a combined feature set of demographic and HFTM-derived variables. This process retained four features, including one demographic variable (patient age) and three features derived from HFTM fever dynamics: fever timing relative to transplant, a Fast Fourier Transform (FFT)-derived feature representing dominant temperature cycle frequency, and a Continuous Wavelet Transform (CWT)-derived feature capturing mid-range temperature fluctuations.

To mitigate potential data leakage arising from the inclusion of multiple fever episodes from the same patient, model training and evaluation were then performed using repeated stratified 5-fold cross-validation at the patient level, ensuring that all fever episodes from a given patient were assigned exclusively to either training or testing sets within each split. This approach approximated an 80:20 training–testing split in each fold and was repeated across 20 iterations, yielding 100 total training–testing combinations. Hyperparameter tuning was performed using grid search for linear models and random search for non-linear models, with the area under the precision–recall curve (AUCPR) prespecified as the primary performance metric due to class imbalance and the clinical need to optimize both model precision and sensitivity. For hyperparameter tuning, 20% of the training data was reserved within each split as a validation subset.

Across the classification models, most demonstrated the ability to distinguish engraftment fevers from other fever etiologies, with a range of performance characteristics (Fig. 2c). Among these, the logistic regression classifier achieved an AUCPR of 0.84 (95% CI 0.69–0.99) and demonstrated a combination of high specificity (0.94; 95% CI, 0.84–1.0), with a corresponding sensitivity for identifying engraftment fevers of 0.41 (95% CI, 0.13–0.69) (Fig. 2c–d). SHAP (SHapley Additive exPlanations) analysis was subsequently performed to evaluate the relative contribution of the four retained features to model predictions (Fig. 2f). While patient age and fever timing relative to transplant demonstrated the largest overall contribution to classification performance, an FFT-derived feature representing dominant temperature cycling behavior and a CWT-derived feature capturing mid-range temperature fluctuations both contributed substantially to model predictions, supporting the importance of dynamic temperature characteristics in distinguishing fever etiology (Fig. 2f, Supp. Fig. S7).

Our results provide a proof-of-concept for the potential of near real-time diagnosis of fever etiology using continuous temperature data collected using non-invasive, wearable technology. In practical terms, our results, correctly identifying roughly half of engraftment fevers (i.e. sensitivity of 0.41) and almost all non-engraftment fevers (i.e. specificity of 0.94), would extrapolate to reducing unnecessary antibiotic exposure in a clinically-meaningful number of HCT recipients, particularly given how frequently engraftment fevers occur^19,20^. In our own fever cohort—where 37% (25 out of 68) febrile episodes were engraftment-related—this model could theoretically have enabled antibiotic de-escalation in approximately 11 of the 25 engraftment cases. Assuming a typical 5-day antibiotic course for culture-negative neutropenic fever, this corresponds to ∼55 antibiotic-days averted, in addition to reductions in unnecessary diagnostic testing and hospital length of stay among the 38 patients whose fever data was used for model training and testing. Furthermore, reducing antibiotic exposure in patients receiving allogeneic HCT might reduce the occurrence of clinically significant GVHD^8,9^. Importantly, the model’s risk of misclassifying antibiotic-requiring fever events as engraftment was low. Among all potential fevers tested across 100 iterations (repeated cross-validation), the model rarely incorrectly labeled clinician adjudicated bacterial infections (either culture positive or negative) as engraftment (median 0.0%, IQR 7.0% across held-out folds); likewise, when tested using only “ground truth” bacterial infection-related fevers, the model had similarly strong discriminatory performance (median 0.0%, IQR 17.0%) (Supp. Fig S8).

The integration of wearable sensors and continuous vital-sign monitoring into care of immunocompromised cancer patients remains an emerging and largely untapped domain, with substantial opportunities to improve real-time assessment of patient status^24,25^. To date, most efforts in this area have centered on the early detection of fever^13,14,26,27^. While early recognition is essential, it is increasingly insufficient on its own; as wearable technologies become more sensitive, distinguishing the underlying etiology of fever becomes critical to avoid overwhelming clinicians with notifications for what could otherwise be benign causes such as engraftment. Our study represents the first demonstration of how continuous temperature data can be systematically processed and analyzed to build predictive models capable of making clinically relevant predictions of fever diagnoses in patients undergoing HCT. This framework provides a foundation not only for differentiating engraftment fevers from other causes, but also in the future could be extended to include continuous physiologic monitoring to detect a broader range of treatment-related toxicities and infectious complications in patients receiving active cancer therapy.

Importantly, this study should be considered a proof-of-concept investigation demonstrating that unique signals captured from continuous HFTM data can inform the diagnosis of fever etiology, rather than establishing a generalizable predictive model. Because the model was trained and evaluated on a relatively small, single-center dataset (n= 68 HFTM traces), it may not fully capture the heterogeneity of engraftment-related temperature patterns, and the findings should not be interpreted as broadly generalizable. We anticipate that future studies in larger patient populations using HFTM data will enable the development of more refined models that better represent under-sampled temperature patterns and improve classification boundaries, potentially confirming generalizability while also improving model sensitivity with preservation of high specificity and precision. Additionally, several fever episodes in this study could not be analyzed due to degraded data quality during periods when patients were acutely ill, often related to inconsistent compliance with sensor placement. As wearable temperature monitoring becomes more routinely integrated into standard clinical care pathways—rather than relying on patient self-application without clinical staff assistance, as was the case in this study—we expect that adherence and data fidelity will be much improved, thereby reducing missing data in future studies.

In summary, this work demonstrates that continuous temperature data collected from patients outfitted with wearable sensors can be leveraged not only for early fever detection but can also inform diagnosing the etiology of fever using machine learning–based analysis. Such an approach offers the potential of a future in which antibiotics could be de-escalated (i.e. early discontinuation) in cases where machine learning models based in part on HFTM data predict engraftment-related or other fevers not needing therapy with antibiotics, providing a novel and actionable tool to strengthen antibiotic stewardship in HCT care. Moving forward, prospective studies will be important for refining model performance through inclusion of additional physiologic and clinical features along with validation in larger sample size patient populations. Together, we envision that these efforts will help establish continuous physiologic monitoring using wearable devices as a scalable platform for real-time diagnostic support in immunocompromised cancer patients who develop fever.

## Resource Availability

Lead Contact: Further information and requests for materials and resources should be directed to the lead contact, Dr. Muneesh Tewari.

Materials Availability: This study did not generate new, unique reagents.

Data and code availability: https://github.com/tewarilab/hftm-fever-patterns-release/. The University of Michigan’s Innovation Partnerships (UMIP) unit will handle potential charges/arrangements of the use of data by external entities, using such methods as data use agreements. Please contact UMIP for inquiries.

## Data Availability

Analyzed data (i.e., results) corresponding to the figures and display items presented in this manuscript are available upon reasonable request to qualified non-commercial entities for non-commercial, scientific research purposes. Primary individual-level participant data may not be shareable publicly due to ethical and privacy restrictions mandated by IRB-approved protocols. Access to the requested analyzed data items is subject to the receipt of necessary institutional approvals, compliance with applicable regulatory frameworks, and the execution of a formal Data Use Agreement (DUA), if required by the University of Michigan. Researchers interested in requesting these data items should contact the corresponding authors.

## Acknowledgements

This work was supported by a Taubman Medical Institute Grand Challenge grant, a Taubman Institute Innovation Project grant, and NIH grant K24HL156896. SNK was supported by an NIH Training Grant (T32 CA009357). CF was supported by an NIH Training Grant (T32 HL007622).

## Declaration of generative AI and AI-assisted technologies in the manuscript preparation process

During the preparation of this work, the author(s) used ChatGPT (OpenAI) and Claude (Anthropic) to assist with language editing and improvement of text clarity and readability. The author(s) reviewed and edited the output as needed and take full responsibility for the content of the published article.

