## Supplementary Information for "Rapid diagnosis of fever etiology using wearable temperature monitoring and machine learning"

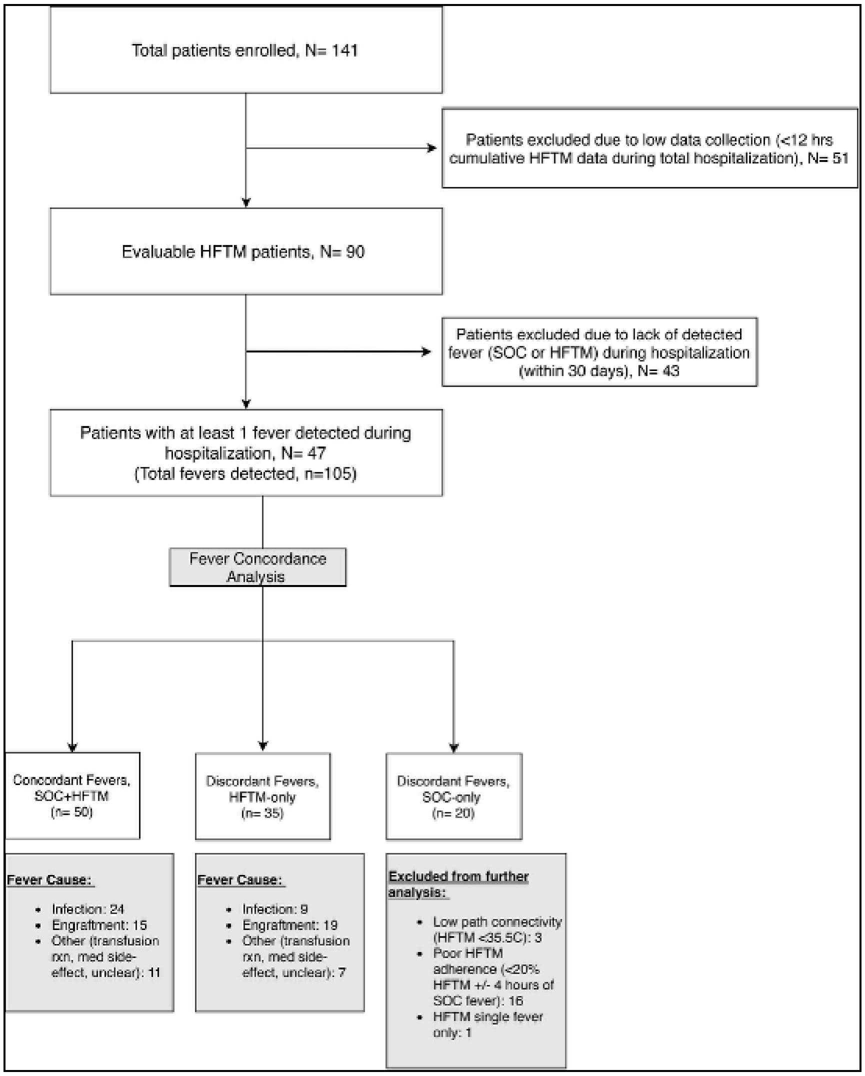


**Supplementary Figure S1. Study enrollment, cohort derivation, and fever concordance analysis.** Flow diagram illustrating patient enrollment, exclusions, and classification of febrile events included in fever concordance analysis. Of 141 enrolled patients, 51 were excluded due to insufficient cumulative HFTM data (<12 hours during entire hospitalization), which was likely due to the fact that the study relied on patients to self-apply and change sensors during their hospitalization. Among the remaining 90 evaluable patients, 43 had no fever detected by either SOC thermometry or HFTM during hospitalization up to 30 days and were excluded. Forty-seven patients experienced at least one febrile episode, yielding 105 total fevers. Febrile events were categorized as concordant (detected by both SOC and HFTM), discordant HFTM-only, or discordant SOC-only. Concordant and HFTM-only fevers were adjudicated by clinical review and classified by presumed etiology (infection, engraftment-related, or other). None of the SOC-only fevers met criteria for downstream analyses comparing them with HFTM because in all cases they were due to either incomplete HFTM data collection, or low signal quality around the time of the SOC fever event, or in one case the SOC-only fever was excluded because there was an associated HFTM fever reading >38.0°C but it did not meet our internal criterion for an HFTM fever (at least three (3)

independent temperature measurements ≥38.0°C within a 1-hour period of the initial ≥38.0°C HFTM measurement).

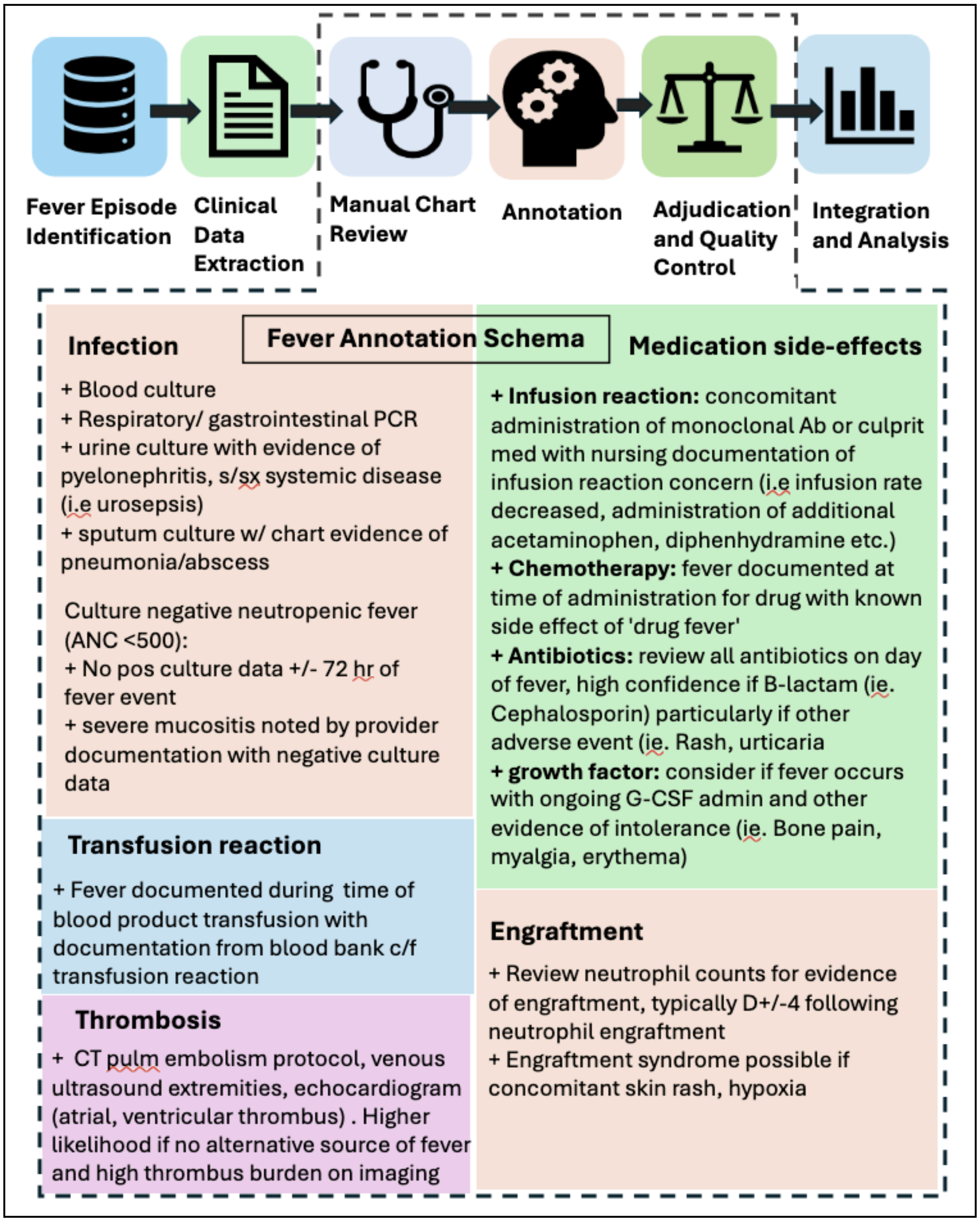


**Supplementary Figure S2. Schematic of fever episode adjudication workflow and annotation schema.** Analytic workflow for initial fever episode identification, clinical data extraction, application of a standardized fever annotation schema, manual chart review, adjudication and quality control, and data integration and analysis. The annotation schema defines criteria used by clinician annotators to adjudicate etiologies for each individual fever episode including infection, medication side-effects, transfusion reaction, thrombosis, and engraftment.

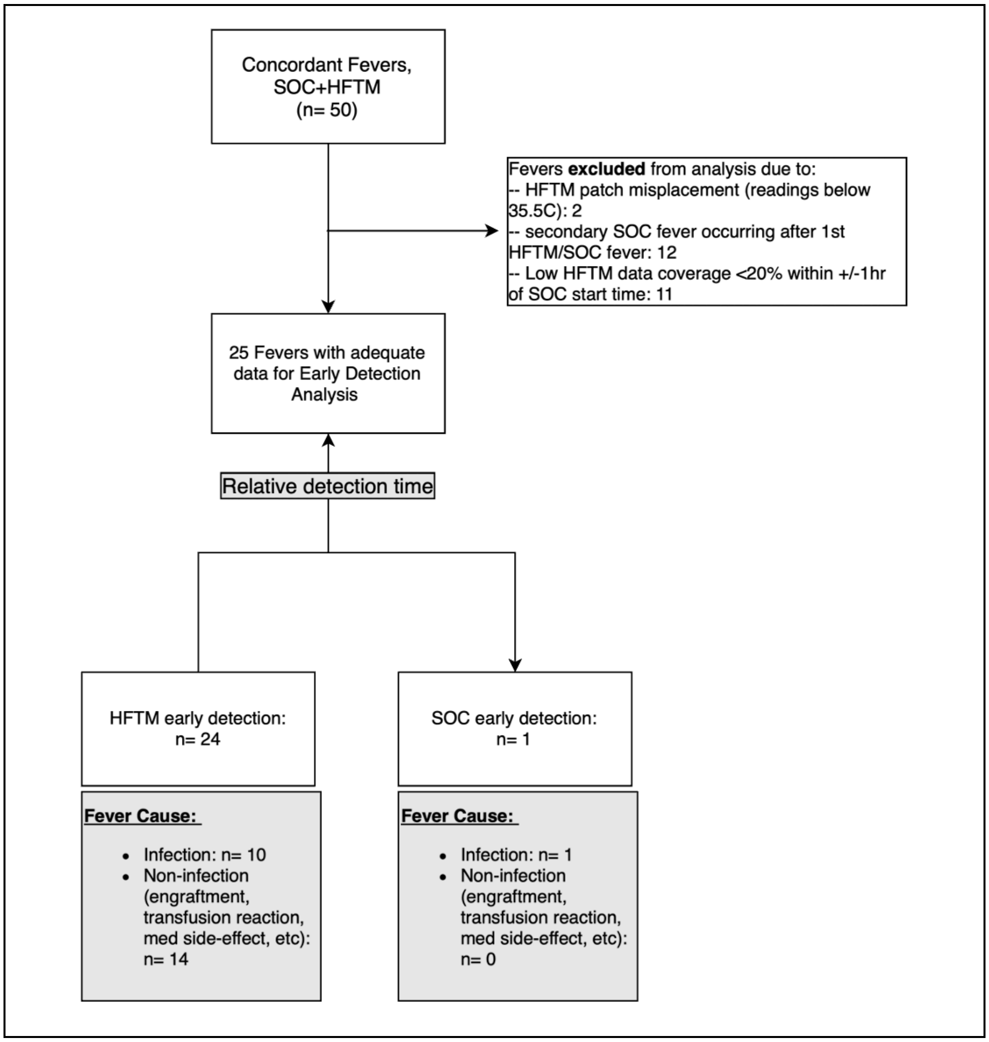


**Supplementary Figure S3. Schema for selecting fever events for early fever detection analysis among concordant fever episodes.** Flow diagram illustrating inclusion, exclusion, and classification of concordant fever episodes (detected by both SOC and HFTM) for the early detection analysis. Among 50 concordant fever episodes, 25 met prespecified data quality and coverage criteria and were included in the analysis. Exclusion criteria included evidence of HFTM patch misplacement (e.g., non-physiologic temperature readings ≤35.5 °C), secondary SOC fevers occurring after an initial concordant episode, and insufficient HFTM data coverage (<20%) within ±1 hour of the SOC fever start time. For included episodes, fever onset times were independently determined for SOC and HFTM, and relative detection time was calculated as the difference between HFTM and SOC fever start times. Early detection was defined as earlier fever onset by HFTM or SOC, with negative values indicating earlier detection by HFTM and positive values indicating earlier detection by SOC. Fever episodes were further stratified by adjudicated etiology (Infection vs. non-infection).

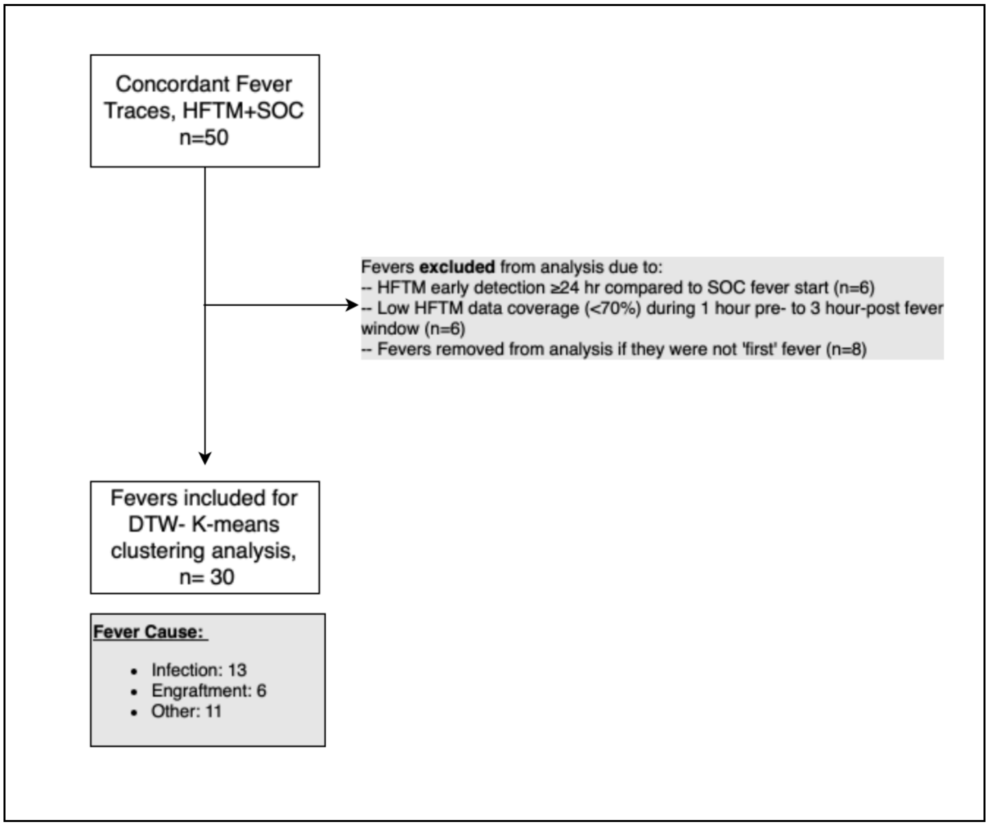


**Supplementary Figure S4. Preprocessing and fever event selection for fever trace clustering analysis.** Flow diagram illustrating selection of concordant fever events for dynamic time warping (DTW)–based clustering analysis. Among 50 concordant fever traces detected by both high-frequency temperature monitoring (HFTM) and standard-of-care thermometry (SOC), events were excluded based on predefined criteria including excessive temporal discordance between HFTM and SOC fever onset (≥24 hours), insufficient HFTM data coverage (<70%) during the peri-fever window (1 hour prior to 3 hours after fever onset), or if the event did not represent the first fever event for that patient during hospitalization. After preprocessing and quality control, 30 fever traces were retained for DTW-based k-means clustering analysis. The distribution of adjudicated fever etiologies among included traces is shown.

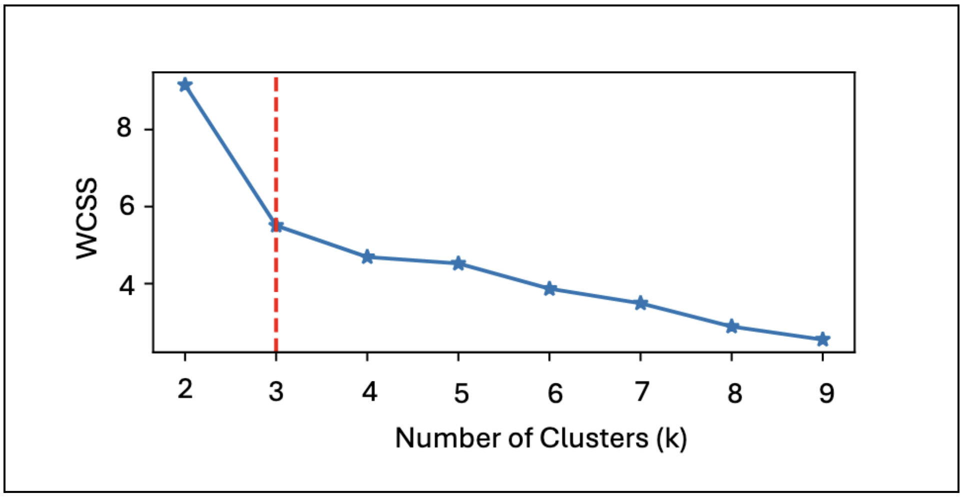


**Supplementary Figure S5. Elbow method for selection of the number of clusters in k-means clustering of temperature traces.** Plot of within-cluster sum of distances (WCSS) across candidate values of *k* for k-means clustering of continuous temperature traces using dynamic time warping (DTW)–based distances. Pairwise DTW distances were calculated and used as the similarity metric for clustering with the TimeSeriesKMeans algorithm ('tslearn', Python package). The elbow point at $k = 3$ (dashed red line) was selected as the optimal number of clusters based on diminishing reductions in within-cluster distances beyond this value, and was used for subsequent clustering and interpretation of fever trace patterns.

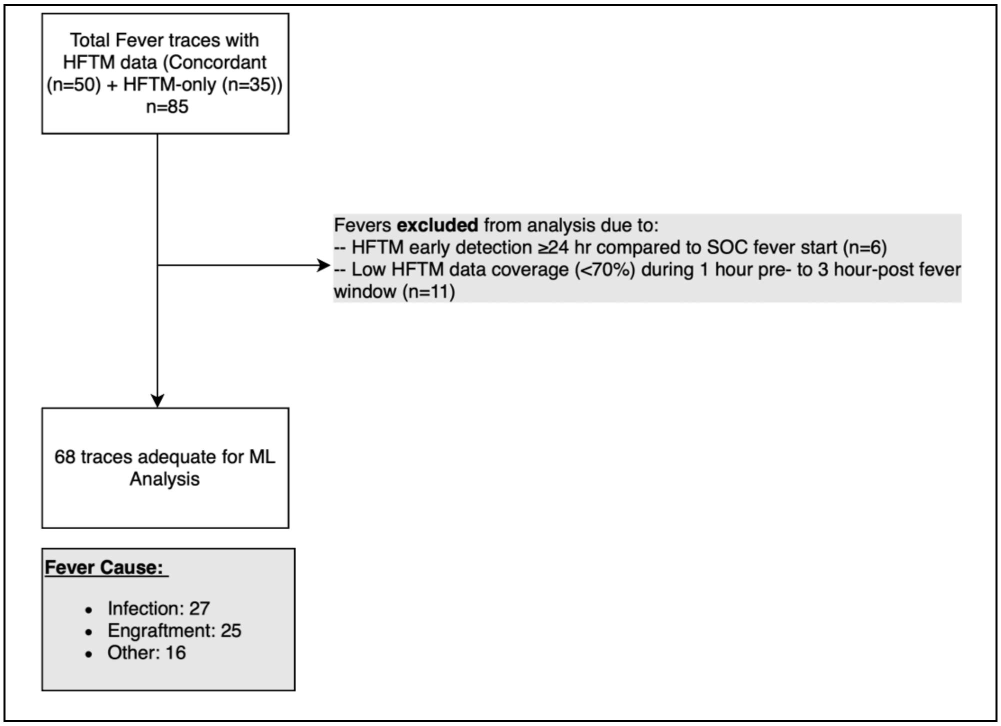


**Supplementary Figure S6. Fever trace preprocessing and fever event selection for machine-learning model training and testing.** Flow diagram illustrating preprocessing, quality control, and inclusion of HFTM fever traces for machine-learning analyses. Temperature traces from concordant fevers (detected by both SOC thermometry and HFTM) and HFTM-only fevers were initially aggregated (n = 85). Prolonged HFTM fever events concordant with multiple SOC fevers were subdivided into distinct fever episodes to ensure one-to-one correspondence between HFTM traces and SOC-defined fever etiologies. Fever traces were excluded based on predefined criteria, including excessive temporal discordance between HFTM and SOC fever onset (≥24 hours) and insufficient HFTM data coverage (<70%) during the peri-fever window (1 hour prior to 3 hours following fever onset). After preprocessing and quality control, 68 fever traces were retained for downstream model development and evaluation and were assigned binary class labels based on clinical adjudication (engraftment vs non-engraftment etiologies).

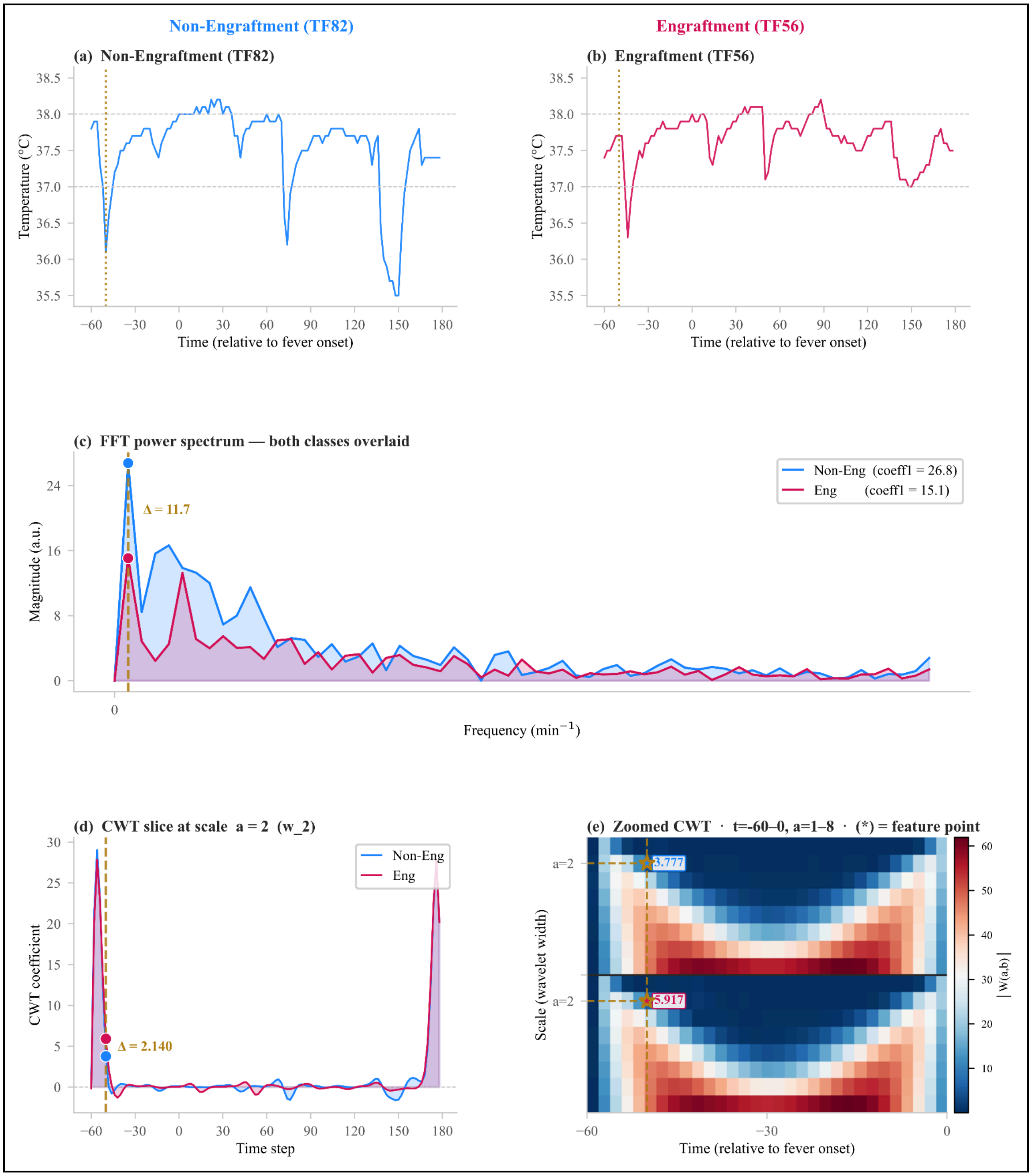


**Supplementary Figure S7. FFT- and CWT-derived features distinguishing engraftment from non-engraftment fevers. (a, b)** Raw wearable temperature traces (°C) for the two example fevers — TF82 (non-engraftment, blue) and TF56 (engraftment, red) — plotted against time relative to fever onset (min). The two horizontal dashed lines mark the 37.0 °C reference and the 38.0 °C fever threshold. The vertical dotted line marks the time point (t = −50 min) corresponding to coefficient index 5, the CWT sample extracted as the feature in panels (d, e) **(c)** FFT magnitude spectra of the two traces, overlaid. Engraftment fevers, driven by a slow, smooth temperature rise, concentrate their energy in the lowest-frequency components. The vertical dashed line marks the frequency bin of the first FFT coefficient (coeff1), and the marker highlights each class's magnitude at that bin (non-engraftment 26.8 vs engraftment 15.1 a.u.). Δ = 11.7 a.u. is the absolute between-class difference in the coeff1 magnitude — the separation that makes this coefficient discriminative. **(d)** Longitudinal slice through the continuous wavelet transform at scale a = 2 (wavelet width 2), i.e., |W(a = 2, b)| as a function of time for each class. The bracket and Δ = 2.140 mark the value extracted as the feature coeff_5__w_2 (coefficient index 5 of this slice) and its between-class difference. **(e)** Zoomed CWT scalograms (|W(a, b)|) for each class over scales a = 1–8 and time t = −60 to 0 min. The asterisk (*) marks the exact coordinate — coefficient index 5, scale a = 2 — that tsfresh extracts as coeff_5__w_2. Engraftment shows higher wavelet activation at this point (5.917) than non-engraftment (3.777); the 2.140 gap between them is the Δ quantified in (d).

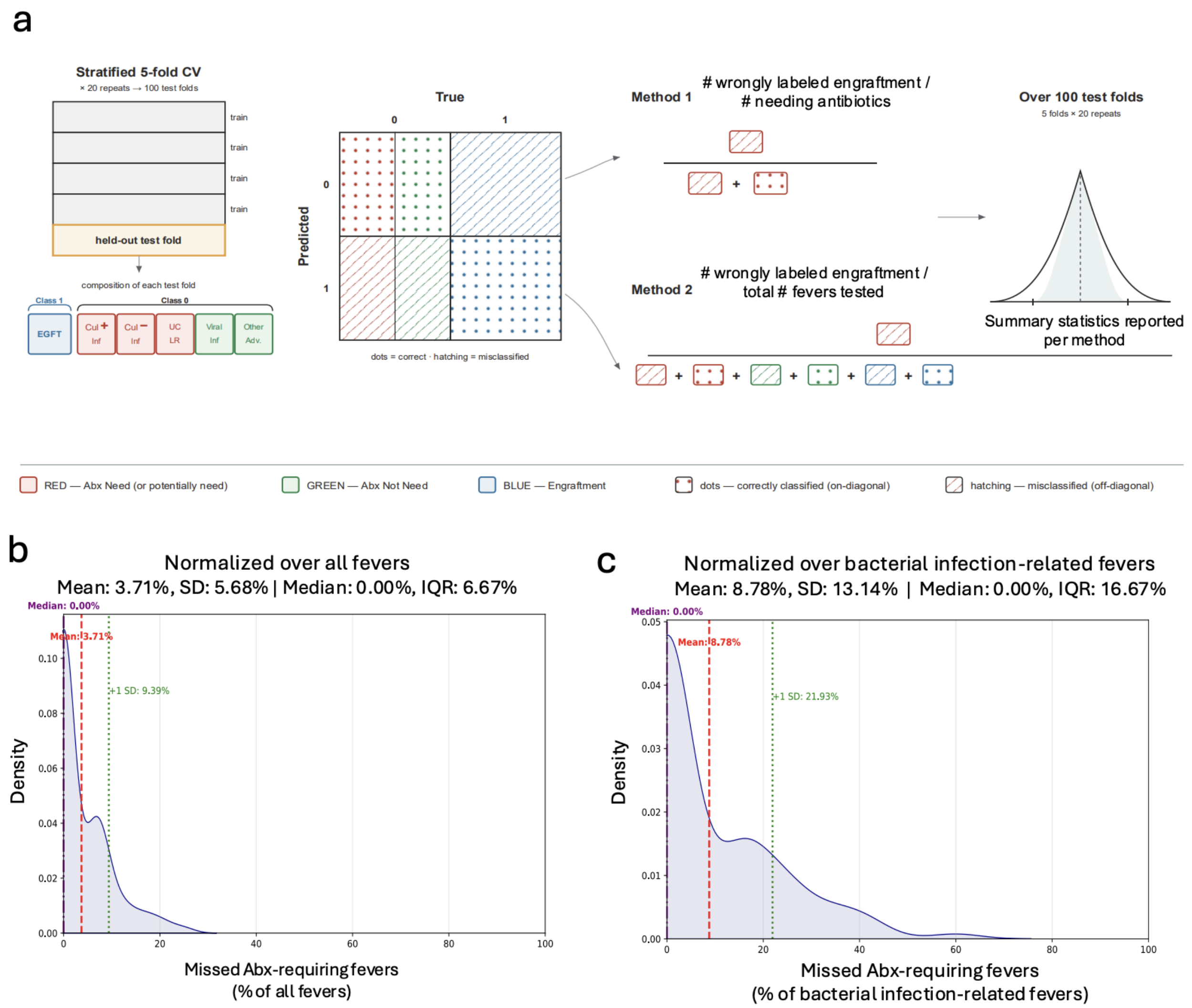


**Supplementary Figure S8. Distribution of the rate at which antibiotic-requiring fevers are misclassified as engraftment across 100 train–test folds.** Kernel density estimates of the fold-level miss rate over the 100 held-out folds (5-fold cross-validation × 20 repeats). **(a)** In every fold, the numerator is the count of fevers that require antibiotics, but are assigned to the engraftment class by the model (ie. false positives). Fever event requiring antibiotics were defined as culture positive bacterial infections (Cul+ inf) and culture negative infections (Cul- inf) that were adjudicated as culture negative neutropenic fever by clinical reviewers. For the purposes of these analyses, we also labelled all "unclear" fever etiologies as antibiotic requiring events. The median (solid purple line), mean (red dashed line), and standard deviation (+1 SD, green dotted line) are shown on plots for both distributions. **(b)** Rate normalized over all fevers encountered in the fold —ie. the fraction of the total fevers tested across cross validation misclassified by the model as engraftment when they were actually antibiotic-requiring fevers. Median: 0.00%, mean: 3.71%, +1 SD: 9.39%. **(c)** Rate normalized over only true antibiotic-requiring infections tested during model cross validation — ie. the fraction of true antibiotic-requiring infections misclassified by the model as engraftment when they were actually antibiotic-requiring infections. Median: 0.00%, mean: 8.78%, +1 SD: 21.93%. The median of 0.00% in both panels indicates that in more than half of the folds no antibiotic-requiring fever was misclassified; the larger mean and wider spread in (b) follow from its smaller, infection-only denominator.

**Supplementary Table 1. Fever annotation summary.** Clinical annotation of all febrile episodes included in the study (n = 105). Each row corresponds to a unique fever episode and includes patient identifier and fever number, method of fever detection, timing of fever based on day post-transplant (DPI), timing relative to neutrophil engraftment, microbiologic findings, concurrent medications of interest, blood product exposure at time of fever, and adjudicated fever etiology. In the patient/fever identifier column, fever detection method is denoted as HFTM-only (T), standard-of-care thermometry only (SOC-only; S), or concordant detection by both methods (C).

*Please note that 19 of the n=20 fevers designated as SOC-only (S) in fact had missing HFTM data at the time of SOC fever, or poor HFTM signal quality, and therefore cannot be reliably considered SOC-only and were excluded for early detection of fever analysis as detailed elsewhere in this article.*

Fever etiologies were assigned by retrospective clinical adjudication based on clinical data extracted from the EHR, including culture results, laboratory values, medication exposures, transfusion timing, and clinical documentation as per the standardized approach described elsewhere in this article. Neutrophil engraftment was defined as the first of three consecutive days with an absolute neutrophil count >0.5 × $10^9$/L. Abbreviations are defined below the table.

| Pt (ID)- Fever# (T/S/C) | DPI | Day of Engraftment | Infection | Notable Medications | Blood Products | Fever Etiology |
|---|---|---|---|---|---|---|
| HCT (1)-Fever#1(T) | D+5 | - | - | - | - | CNFN |
| HCT (2)-Fever#1(T) | D+8 | - | BCN | MGF | - | CNFN |
| HCT (2)-Fever#2(S) | D+9 | - | BCN | MGF | - | CNFN |
| HCT (4)-Fever#1(T) | D+1 | - | ACN | BSA: VA/AZT | - | CNFN |
| HCT (5)-Fever#1(C) | D+2 | - | ACN | BSA: FEP | none | CNFN |
| HCT (5)-Fever#2(T) | D+13 | - | - | - | - | CNFN |
| HCT (6)-Fever#1(S) | D+2 | - | BCP: CLABSI | BSA: VA/FEP MGF | - | MDI-B |
| HCT (7)-Fever#1(T) | D+9 | D+12 | BCN | BSA: VA | - | NE |
| HCT (11)-Fever#1(S) | D+4 | - | - | - | PRBC | ATR |
| HCT (11)-Fever#2(C) | D+9 | - | ACN | MGF | - | CNFN |
| HCT (12)-Fever#1(C) | D+5 | - | BCP: S. mitis | BSA: VA/FEP | - | MDI-B |

| | | | | | | |
|---|---|---|---|---|---|---|
| HCT (12)-Fever#2(C) | D+8 | - | RPP (+) | BSA: FEP | - | MDI-V |
| HCT (12)-Fever#3(C) | D+15 | D+14 | ACN | BSA: FEP MGF | - | NE |
| HCT (13)-Fever#1(T) | D+9 | D+12 | ACN | MGF | - | NE |
| HCT (13)-Fever#2(T) | D+12 | D+12 | ACN | MGF | - | NE |
| HCT (14)-Fever#1(C) | D+6 | - | ACN | - | - | Hemolysis PLS |
| HCT (14)-Fever#2(C) | D+8 | - | ACN | - | - | Hemolysis PLS |
| HCT (16)-Fever#1(C) | D-12 | - | ACN | ATG | - | IR |
| HCT (16)-Fever#2(C) | D+6 | - | ACN | BSA: FEP MGF | - | CNFN |
| HCT (20)-Fever#1(C) | D+0 | | BCP: MAC | - | - | MDI-B |
| HCT (22)-Fever#1(S) | D-4 | - | ACN | ATG | - | IR |
| HCT (22)-Fever#2(C) | D-2 | - | ACN | ATG | - | IR |
| HCT (22)-Fever#3(T) | D+14 | - | BCP: S. epi | BSA: FEP/VA MGF | - | MDI-B |
| HCT (22)-Fever#4(C) | D+15 | - | BCP: S. epi | BSA: FEP | - | MDI-B |
| HCT (23)-Fever#1(T) | D-3 | - | ACN | FluBu | - | CNFN |
| HCT (23)-Fever#2(C) | D-1 | - | ACN | - | - | HLH |
| HCT (23)-Fever#3(T) | D+6 | - | BCP: CLBSI E. faecium | BSA: FEP | - | MDI-B |
| HCT (23)-Fever#4(C) | D+8 | - | BCP: CLBSI E. faecium | BSA: FEP | - | MDI-B |
| HCT (23)-Fever#5(C) | D+11 | D+13 | ACN | BSA: FEP/VA | - | NE |
| HCT (23)-Fever#6(C) | D+12 | D+13 | ACN | BSA: FEP/VA | - | NE |
| HCT (23)-Fever#7(C) | D+15 | D+13 | ACN | BSA: FEP/VA | - | NE |
| HCT (23)-Fever#8(T) | D+18 | D+13 | - | BSA: FEP/VA | - | NE |

| | | | | | | |
|---|---|---|---|---|---|---|
| HCT (23)-Fever#9(T) | D+22 | - | - | MGF | - | DIF |
| HCT (24)-Fever#1(S) | D+6 | - | ACN | BSA: FEP/VA | - | MDI-B |
| HCT (24)-Fever#2(C) | D+7 | - | ACN | BSA: FEP/VA | - | MDI-B |
| HCT (25)-Fever#1(C) | D+1 | - | ACN | BSA: FEP | - | Unclear |
| HCT (25)-Fever#2(T) | D+3 | - | RPP (+) Rhino-enterovirus | BSA: FEP/VA | - | MDI-V |
| HCT (25)-Fever#3(C) | D+5 | - | RPP (+) Rhino-enterovirus | zosyn | - | MDI-V |
| HCT (25)-Fever#4(C) | D+8 | - | RPP (+) Rhino-enterovirus | BSA: FEP/VA | - | MDI-V |
| HCT (27)-Fever#1(T) | D+8 | - | - | MGF | - | DIF |
| HCT (27)-Fever#2(C) | D+10 | - | ACN | BSA: FEP MGF | - | CNFN |
| HCT (27)-Fever#3(C) | D+13 | - | ACN | BSA: FEP | - | CNFN |
| HCT (27)-Fever#4(C) | D+15 | - | Lung biopsy aspergillosis (+) | BSA: FEP | - | MDI-F |
| HCT (27)-Fever#5(C) | D+17 | - | Lung biopsy aspergillosis (+) | BSA: FEP/Mica/Vori | - | MDI-F |
| HCT (27)-Fever#6(C) | D+21 | - | Lung biopsy aspergillosis (+) | BSA: FEP/Mica/Vori | - | MDI-F |
| HCT (31)-Fever#1(S) | D+2 | - | ACN | - | - | No HFTM data collected |
| HCT (31)-Fever#2(T) | D+13 | - | BCP: S. epi, R. mucilaginosa | BSA: VA | - | MDI-B |
| HCT (31)-Fever#3(S) | D+14 | - | BCP: S. epi, R. mucilaginosa | BSA: FEP/VA MGF | - | MDI-B |
| HCT (32)-Fever#1(T) | D+6 | - | - | - | - | CNFN |
| HCT (32)-Fever#2(S) | D+9 | D+12 | ACN | - | - | NE |

| | | | | | | |
|---|---|---|---|---|---|---|
| HCT (34)-Fever#1(C) | D+9 | D+11 | ACN | BSA: FEP MGF | - | NE |
| HCT (34)-Fever#2(C) | D+10 | D+11 | ACN | BSA: FEP MGF | - | NE |
| HCT (36)-Fever#1(C) | D+10 | D+12 | ACN | MGF | - | NE |
| HCT (38)-Fever#1(S) | D-3 | - | ACN | BSA: FEP | - | Unclear |
| HCT (39)-Fever#1(T) | D+9 | D+11 | - | MGF | - | NE |
| HCT (40)-Fever#1(S) | D+9 | - | RPP (+) Parainfluenza | - | - | MDI-V |
| HCT (42)-Fever#1(T) | D+11 | D+13 | none | MGF | - | NE |
| HCT (49)-Fever#1(T) | D+11 | D+12 | - | MGF | - | NE |
| HCT (50)-Fever#1(C) | D+6 | D+12 | ACN | MGF | - | NE |
| HCT (50)-Fever#2(C) | D+8 | D+12 | ACN | MGF | - | NE |
| HCT (51)-Fever#1(C) | D+5 | - | BCP: E. coli | BSA: meropenem | - | MDI-B |
| HCT (52)-Fever#1(C) | D+7 | D+13 | ACN | MGF | - | NE |
| HCT (52)-Fever#2(C) | D+8 | D+13 | ACN | MGF | - | NE |
| HCT (52)-Fever#3(T) | D+13 | D+13 | - | MGF | - | NE |
| HCT (53)-Fever#1(S) | D+9 | NA | NA | NA | NA | No HFTM data collected |
| HCT (53)-Fever#2(T) | D+10 | D+12 | - | MGF | - | NE |
| HCT (54)-Fever#1(T) | D+7 | D+13 | - | - | - | NE |
| HCT (58)-Fever#1(C) | D+2 | - | ACN | BSA: FEP | - | CNFN |
| HCT (58)-Fever#2(C) | D+6 | - | RPP (+) COVID19 | BSA: Zosyn | - | MDI-V |
| HCT (58)-Fever#3(C) | D+7 | - | RPP (+) COVID19 | BSA: Zosyn | - | MDI-V |
| HCT (61)-Fever#1(T) | D+9 | D+12 | - | MGF | - | NE |
| HCT (62)-Fever#1(C) | D-14 | - | ACN | Alemtuzuma b | - | IR |
| HCT (62)-Fever#2(C) | D-7 | - | ACN | Fludarabine | - | Unclear |

| | | | | | | |
|---|---|---|---|---|---|---|
| HCT (65)-Fever#1(C) | D+7 | - | ACN | BSA: FEP MGF | - | Unclear |
| HCT (65)-Fever#2(T) | D+11 | - | ACN | BSA: FEP MGF | - | CNFN |
| HCT (65)-Fever#3(T) | D+23 | D+24 | ACN | MGF | - | NE |
| HCT (65)-Fever#4(T) | D+25 | D+24 | ACN | MGF | - | NE |
| HCT (66)-Fever#1(T) | D+7 | D+12 | ACN | MGF | - | NE |
| HCT (66)-Fever#2(T) | D+10 | D+12 | ACN | MGF | - | NE |
| HCT (71)-Fever#1(S) | D+9 | NA | NA | NA | NA | Excluded: No HFTM data collected |
| HCT (71)-Fever#2(S) | D+11 | NA | NA | NA | NA | Excluded: No HFTM data collected |
| HCT (71)-Fever#3(S) | D+13 | NA | NA | NA | NA | Excluded: No HFTM data collected |
| HCT (73)-Fever#1(C) | D+9 | D+12 | ACN | BSA: Zosyn MGF | - | NE |
| HCT (73)-Fever#2(T) | D+13 | D+12 | ACN | BSA: Zosyn MGF | - | NE |
| HCT (74)-Fever#1(C) | D-1 | - | BCP: E. coli | BSA: FEP | - | MDI-B |
| HCT (74)-Fever#2(C) | D+5 | D+8 | ACN | BSA: FEP | - | NE |
| HCT (77)-Fever#1(C) | D+9 | D+12 | ACN | BSA: FEP/VA MGF, HDC | - | ES* |
| HCT (77)-Fever#2(T) | D+16 | - | ACN | BSA: FEP | none | Unclear |
| HCT (77)-Fever#3(C) | D+19 | - | ACN | BSA: FEP | none | Unclear |
| HCT (79)-Fever#1(S) | D+2 | NA | NA | NA | NA | Excluded: No HFTM data collected |
| HCT (79)-Fever#2(S) | D+4 | NA | NA | NA | NA | Excluded: No HFTM data collected |
| HCT (79)-Fever#3(S) | D+5 | NA | NA | NA | NA | Excluded: No HFTM data collected |

| | | | | | | |
|---|---|---|---|---|---|---|
| HCT (82)-Fever#1(C) | D+4 | D+10 | ACN | BSA: FEP | - | NE |
| HCT (83)-Fever#1(C) | D+8 | D+10 | ACN | BSA: FEP | - | NE |
| HCT (84)-Fever#1(C) | D+2 | - | BCP: S. mitis CLABSI | BSA: FEP, flagyl, tobramycin | - | MDI-B |
| HCT (84)-Fever#2(C) | D+7 | - | BCP: S. mitis CLABSI | BSA: FEP, flagyl, tobramycin | - | MDI-B |
| HCT (84)-Fever#3(T) | D+8 | D+12 | ACN | BSA: FEP | - | NE |
| HCT (84)-Fever#4(S) | D+11 | NA | NA | NA | NA | Excluded: No HFTM data collected |
| HCT (84)-Fever#5(S) | D+14 | NA | NA | NA | NA | Excluded: No HFTM data collected |
| HCT (86)-Fever#1(T) | D+7 | D+10 | ACN | MGF | - | NE |
| HCT (91)-Fever#1(T) | D+4 | - | - | - | - | Unclear |
| HCT (91)-Fever#2(T) | D+11 | D+16 | ACN | MGF | - | NE |
| HCT (91)-Fever#3(T) | D+13 | D+16 | ACN | MGF | - | NE |
| HCT (95)-Fever#1(S) | D+0 | NA | NA | NA | NA | Excluded: No HFTM data collected |
| HCT (95)-Fever#2(C) | D+5 | - | BCP: Respiratory Cx (+) MRSA | BSA: Zosyn ,VA | - | MDI-B |

Abbreviations: culture negative febrile neutropenia (CNFN); neutrophil engraftment (NE); blood cultures negative (BCN); blood cultures positive (BCP); all cultures (e.g blood, urine, respiratory) negative (ACN); Broad-spectrum antibiotics (BSA); Vancomycin (VA); Cefepime (FEP); central-line associated blood stream infection (CLABSI); drug-induced fever (DIF); myeloid growth factor (e.g granulocyte colony-stimulating factor) (MGF); microbiologically defined infection (MDI), MDI-B (Bacterial), MDI-V (Viral), MDI-F (Fungal); acute transfusion reaction (ATR) occurring during infusion packed red blood cell (PRBC) or platelets (PLT); respiratory pathogen panel (RPP); passenger lymphocyte syndrome (PLS); Anti-thymocyte globulin (ATG); infusion reaction (IR); Mycobacterium Avium complex infection (MAC); Hemophagocytic lymphohistiocytosis (HLH); High-dose corticosteroids (HDC); Engraftment syndrome (ES), *One fever event (HCT (77)- Fever#1) was classified as engraftment syndrome (ES)-related rather than neutrophil engraftment (NE)-related. This distinction was made because the patient developed clinically significant illness requiring high-dose corticosteroids, in contrast to NE-associated fevers, which are otherwise benign and do not require steroid therapy.

**Day of Engraftment: “Day+X”, the first of three consecutive days when ANC is greater than $0.5x10^9$/L

**Supplementary Table 2. Patient demographics for those experiencing at least one fever episode during their hospitalization (see Supp. Fig. S1)**

| Variable | Category | Overall |
|---|---|---|
| **Age, mean (SD)** | | 45.0 (21.1) |
| **Race, n (%)** | Caucasian | 40 (85.1) |
| | African American | 3 (6.4) |
| | Asian | 3 (6.4) |
| | Other | 1 (2.1) |
| **Ethnicity, n (%)** | Non-Hispanic or Latino | 44 (93.6) |
| | Hispanic or Latino | 3 (6.4) |
| **Diagnosis, n (%)** | Acute myeloid leukemia | 13 (27.7) |
| | B-ALL | 6 (12.8) |
| | myelodyslastic syndrome | 6 (12.8) |
| | DLBCL | 3 (6.4) |
| | Peripheral T-cell lymphoma | 3 (6.4) |
| | myelofibrosis | 3 (6.4) |
| | aplastic anemia | 2 (4.3) |
| | multiple myeloma | 2 (4.3) |
| | Adult-onset X-linked Cerebral Adrenoleukodystrophy | 1 (2.1) |
| | CMML/MDS | 1 (2.1) |
| | CNS HLH Perforin Deficiency | 1 (2.1) |
| | Chronic Granulomatous Disease | 1 (2.1) |
| | Mantle cell lymphoma | 1 (2.1) |
| | NEMO deficiency | 1 (2.1) |
| | Primary mediastinal B-cell lymphoma with secondary CNS lymphoma | 1 (2.1) |
| | T-ALL | 1 (2.1) |
| | XIAP deficiency | 1 (2.1) |
| **Total** | | 47 |

### Experimental Procedures

**Patient enrollment and data collection.** We conducted an prospective observational study of cancer patients (n=141) outfitted with FDA-approved, non-invasive, axillary temperature patches (TempTraq ®, BlueSpark Technologies) continuously measuring temperature every two minutes (high-frequency temperature monitoring; HFTM) in addition to being monitored by standard-of-care (SOC) nursing vitals during inpatient hematopoietic stem cell transplantation (HCT). After providing institutional review board (IRB)-approved informed consent, patients were instructed to self-administer HFTM patches that were worn daily and replaced every twenty-four hours per the manufacturer's guidelines throughout the duration of their respective hospitalization. HFTM data was transmitted from the TempTraq® axillary patch to a bluetooth gateway device provided by BlueSpark Technologies and subsequently uploaded to a BlueSpark Technologies server. HFTM data was downloaded from the secure server via a mobile application (TempTraq® Clinician) and stored on an encrypted laptop for further analysis. Subject demographic data, nursing SOC recorded temperatures, and dates of admission, stem-cell transplantation, and discharge from hospital was obtained from the electronic health record (EHR) through our institution's 'Research Data Warehouse' and, likewise, stored on an encrypted laptop.

**Data preprocessing.** Patient-matched HFTM and EHR data (temperature recordings and hospital admission dates) for each subject were uploaded into Python (ver. 3.11.5) and RStudio (ver. 1.2.1335; CRAN ver. R-4.0.3). The "HFTM monitoring period" was defined as the time of the first patch application to the last HFTM temperature recording while a patient was hospitalized and only HFTM and clinical-SOC temperature recordings obtained during the monitoring period were evaluated for all downstream analysis. Dates and times were converted to an appropriate format (yyyy:mm:dd hh:mm:ss) and were masked as 'days post-infusion' relative to each individual subject's treatment infusion date. To identify patients with poor patch adherence, those with less than 12 hours of cumulative HFTM data collected during their entire hospitalization were excluded from further analysis. HFTM temperature recordings below 35.5C were discarded as these were presumed to be non-physiologic and due to technical issues (e.g., temperature sensor migration on the skin, away from the axilla).

**Concordance analysis of fevers identified by HFTM and SOC.** To evaluate agreement between high-frequency temperature monitoring (HFTM) and standard-of-care (SOC) temperature measurements in detecting fever, we conducted a concordance analysis using specific event definitions and exclusion criteria. A fever detected by HFTM was defined as at least three (3) independent temperature measurements ≥38.0°C within a 1-hour period of the initial ≥38.0°C HFTM measurement. Fever onset was defined as the time of the first ≥38.0°C HFTM measurement; fever resolution was defined as the time of the last ≥38.0°C HFTM measurement after which no further ≥38.0°C measurements occurred within 24 hours. Any HFTM fever episode lasting less than 1 hour was excluded from analysis.

A fever detected by SOC was defined as a single nursing-recorded temperature ≥38.0°C documented in the EHR. For SOC-detected fevers, fever onset was the time of the first qualifying measurement, and fever resolution was the last ≥38.0°C SOC measurement with no subsequent ≥38.0°C measurement within 24 hours.

Fevers were classified according to the following criteria:

- **Concordant Fever:** A fever detected in the SOC data stream (the "reference" fever) with an HFTM fever detected within 24 hours before or after the start or end time of the reference SOC fever.
- **Discordant—HFTM-only Fever:** A reference fever was detected by HFTM (as defined above), and no fever was detected in the parallel SOC data stream within ±24 hours of the start or end of the HFTM fever episode. HFTM fevers with a duration of less than 1 hour were excluded.
- **Discordant—SOC-only Fever:** A reference fever detected by SOC, with no fever detected in the parallel HFTM data stream within ±24 hours of the start or end of the reference SOC fever.

Certain SOC-only fevers were excluded from concordance analysis and instead reported as "SOC-only due to inadequate HFTM data collection" if HFTM data coverage was less than 20% within ±1 hours of the SOC fever start time. Additionally, if HFTM data demonstrated non-physiological temperature values (e.g., <35.5°C) within the SOC fever window—suggesting TempTraq® patch misplacement—the SOC fever was excluded and reported as "SOC-only due to patch misplacement."

**Clinical annotation of febrile events.** We conducted a retrospective review of the EHR to identify significant clinical events associated with each febrile event detected by HFTM or clinical-SOC. These included evaluation for therapy-related toxicity, sources of infection, medication side-effects, blood product transfusion reactions, and thrombotic events (Supp. Fig. 1). Fever episodes were reviewed and adjudicated by at least two clinician reviewers.

infection fever was considered to be of "high confidence" if positive culture data was present within three days before or after a documented febrile event. Positive culture data was defined as positive blood culture with speciation along with provider team initiation of antibiotics. Respiratory PCR or gastrointestinal PCR data positive for viral or bacterial pathogens commonly associated with fever that were detected within three days of a fever were also considered to be "high confidence." In the case of stool studies positive for c. difficile, the EHR was reviewed for evidence of fulminant disease in order for c. difficile infection to be considered a likely cause of fever. Similarly, in the case of positive urine culture data, we considered urinary sources of fever to be more likely if there was concomitant evidence of pyelonephritis documented by the inpatient provider team or on imaging studies, or evidence of systemic illness indicative of urosepsis. For cases of culture negative fever, we still considered infection to be a possible cause of fever occurring in neutropenic patients (ANC less than 500 or down trending from 1000 in the setting of active cancer treatment), particularly if review of EHR revealed that empiric antibiotics were started by the inpatient provider team or in the case that severe mucositis was documented by the provider time with either initiation of total parenteral nutrition or a patient-controlled analgesia pump.

We considered engraftment fever to be of "strong likelihood" if the first occurrence of fever occurred within 4-days of neutrophil engraftment on patient bloodwork, which was defined as an absolute neutrophil count (ANC) greater than 500 for three consecutive days.

Medication side-effects were also considered as a possible etiology for fever. Infusion reactions were considered "strong likelihood" for fevers occurring with concomitant administration of monoclonal antibodies (i.e. rituximab, etc.) with nursing or provider documentation in the EHR suggestive of infusion reaction concerns as demonstrated by active decision to decrease the infusion rate during infusion, additional administration of tylenol or benadryl or other changes to normal infusion protocol. Chemotherapy was considered a possible etiology if fever was documented within 24 hours of administration, particularly in the case of conditioning chemotherapy regimens. Antibiotic-related fever was considered in the case of persistent, culture-negative fevers, with higher likelihood attributed to beta-lactams such as penicillins or cephalosporins particularly in the case of other documented adverse reactions attributed to antibiotics administration such as urticaria or rash. Finally, fevers occurring secondary to ongoing growth factor administration were considered "strong likelihood" if provider documentation indicated other evidence of growth factor intolerance (i.e bone pain, erythema, or myalgias) necessitating intervention.

As in the case of infusion reaction, we also considered blood product transfusion reactions to be "strong likelihood" in the case of fevers occurring during the time of blood product administration with documentation in the EHR from our institution's blood bank indicating concern for transfusion reaction. Additionally, fevers caused by thrombotic events was considered if no alternative sources of fever could be identified with evidence of high clot burden on confirmatory imaging obtained within 5 days of a fever episode, with acceptable imaging modalities including CT pulmonary embolism protocol, lower extremity doppler studies, and transthoracic echocardiography demonstrating either atrial or ventricular thrombus.

**Early fever detection analysis for concordant events.** Early detection analysis was performed to compare the timing of fever onset during concordant fever episodes. For each concordant fever episode, the fever start time was determined separately for HFTM and SOC according to their respective fever onset definitions. The time difference between the HFTM fever start and the SOC fever start was calculated with a positive value indicating earlier detection by SOC, and a negative value indicating earlier detection by HFTM. To be included in the early fever detection analysis, concordant fever episodes required greater than 20% HFTM data coverage within ±1 hour of the SOC fever start time. Fevers were excluded if this criterion was not met or if there was evidence of patch misplacement (e.g., non-physiologic temperature measurements ≤ 35.5°C).

For each concordant fever episode meeting inclusion criteria, the difference in fever start time between HFTM and SOC was calculated as described above. Early detection times for all eligible episodes were compiled, with negative values indicating earlier detection by HFTM and positive values indicating earlier detection by SOC. To summarize the central tendency of fever detection differences, the median early detection time and corresponding interquartile range (IQR) were calculated. The median and IQR were chosen due to the potential for non-normal distribution of detection time differences. All analyses were performed using the Python statistical package.

**Fever Trace Clustering Using Dynamic Time Warping (DTW):**

**Data preprocessing.** Temperature readings were collected using TempTraq sensors and preprocessed prior to clustering analysis. For each concordant fever event, temperature values were extracted from a time window beginning 1 hour before and extending to 3 hours after the recorded fever onset. Temperature traces containing less than 70% observed (non-missing) data within this window were excluded from further analysis. For the remaining traces, residual missing values were imputed using a two-step approach. Spline interpolation was applied with a window size of 3 and a smoothing factor of 1.8 when sufficient neighboring data points were available. When fewer than three preceding temperature measurements were present, k-nearest neighbor interpolation was used instead. Following preprocessing and quality filtering, a total of 30 high-fidelity temperature traces were retained for downstream clustering analyses.

**DTW and K-means Clustering of HFTM Traces.** Pairwise similarity between preprocessed temperature traces was quantified using dynamic time warping (DTW) (Li, et al. Physiol Meas. 2012), a method previously validated for comparing physiological time-series data. The resulting pairwise DTW distances were used as the similarity metric for clustering. Temperature traces were clustered using the TimeSeriesKMeans algorithm from the tslearn Python package with pairwise DTW distances as the distance metric. The optimal number of clusters was determined using the ‘elbow method’ applied to the within-cluster sum of distances, yielding three clusters ($k = 3$) for subsequent interpretation of fever trace patterns.

**Classification Model Training and Evaluation:**

**Data Preprocessing.** Classification model development was performed using 68 fever episodes derived from HFTM data. Both concordant and HFTM-only fevers were eligible for inclusion. Because the HFTM fever definition considered fever episodes continuous until no additional HFTM fever readings occurred within 24 hours of the preceding fever measurement, some HFTM fever traces spanned prolonged periods and encompassed multiple standard-of-care (SOC) fever events. To ensure that individual fever episodes corresponded to clinically meaningful events, prolonged HFTM traces were manually reviewed and subdivided when multiple SOC fevers occurring within the same HFTM trace were determined to represent clinically independent fever episodes. Fever episodes were considered clinically independent when SOC fever occurrences were separated by more than 24 hours and/or when retrospective clinical adjudication supported distinct underlying fever etiologies. For each additional SOC fever (beginning with the second), we retrospectively examined the preceding 24 hours of HFTM data, assigned a new fever start time using the HFTM fever definition, and defined the end time of the preceding episode as the temperature measurement immediately preceding the subsequent HFTM fever event. This process established a one-to-one mapping between individual fever episodes and their assigned clinical etiologies. Fevers were labeled as a binary outcome based on prior clinical annotation: Class 1 (engraftment fever) versus Class 0 (all non-engraftment fevers, including bacterial infection, culture-negative neutropenia, viral infection, drug-related fever, and other etiologies).

**Feature Extraction and Selection.** Prior to feature extraction, temperature traces — including those from discordant fever events not included in the DTW analysis — were preprocessed using the same interpolation pipeline (spline interpolation with k-nearest neighbor fallback; see section 'Fever Trace Clustering Using Dynamic Time Warping' above). Time-series features were then extracted from these traces using tsfresh and combined with demographic variables. Feature selection was performed using the Boruta algorithm with a Random Forest estimator to identify variables most relevant for distinguishing engraftment from non-engraftment fevers. Class imbalance was addressed by computing an imbalance ratio (IR = number of Class 0 instances / number of Class 1 instances) applied as a class weight. To obtain stable features, Boruta was run on the full dataset 100 times with bootstrap resampling, sampling 80% of fever episodes per run; features selected in at least 20% of runs were retained.

**Model Training and Evaluation.** To prioritize sensitivity to infection-related fevers, etiology-based sample weights were applied during model training: confirmed bacterial infection (weight = 10), high-risk infectious syndromes such as culture-negative neutropenic fever or severe mucositis (weight = 5), and all other etiologies (weight = 1). These weights were applied in addition to the class imbalance weights described above. Model training and evaluation were performed using repeated stratified 5-fold cross-validation with patient-level grouping to prevent information leakage arising from the inclusion of multiple fever episodes from the same patient. This approach yielded approximately 80:20 training-to-testing splits within each fold and was repeated across 20 iterations, resulting in 100 total train-test evaluations. Because patient-level splitting occasionally introduced class imbalance within the training folds, oversampling was applied to approximately restore the original class distribution (25 engraftment versus 43 non-engraftment fevers) prior to model training. We trained and evaluated a suite of classifiers including Logistic Regression, Decision Tree, Random Forest, Naïve Bayes, linear SVM, and XGBoost. Hyperparameters were optimized using grid search for linear models and random search for non-linear models. Model performance was assessed primarily using the area under the precision-recall curve (AUCPR) evaluated on held-out test data.
